# Algorithm Refinement of the National Early Warning Score 2 (NEWS2): Study Protocol

**DOI:** 10.1101/2024.12.10.24318795

**Authors:** Chris Plummer, Madison Milne-Ives, Cen Cong, Lynsey Threlfall, Edward Meinert

**Affiliations:** The Newcastle Upon Tyne Hospitals NHS Foundation Trust, Newcastle upon Tyne, NE7 7DN, UK; NIHR Newcastle Biomedical Research Centre, Newcastle University, Newcastle upon Tyne, NE1 7RU, UK; Translational and Clinical Research Institute, Newcastle University, Newcastle upon Tyne, UK; Department of Primary Care and Public Health, Imperial College London, London, UK; Centre for Health Technology, School of Nursing and Midwifery, University of Plymouth, Plymouth, UK

**Keywords:** National Early Warning Score, clinical deterioration, performance, proof-of-concept

## Abstract

**Introduction:** The second iteration of the National Early Warning Score (NEWS2) has been widely adopted for predicting patient deterioration in healthcare settings using routinely collected physiological observations. The use of NEWS2 has been shown to reduce in-hospital mortality, but it has limited accuracy in the prediction of clinically important outcomes, especially over longer time periods. The increasing implementation of digital patient observations and health records presents an opportunity to investigate whether the addition of individual patient characteristics and information about their care-setting, would improve the predictive accuracy of the score.

**Methods and analysis:** This protocol describes the work to determine whether the performance of the current NEWS2 system could be improved by the use of additional variables. The project has been designed after an extensive scoping review of existing literature on NEWS2 and an exploration of retrospective cohort data in The Newcastle upon Tyne Hospitals NHS Foundation Trust, with input from key clinical stakeholders.

**Ethics and dissemination:** The project has received competitive funding following peer-review, from the NIHR Newcastle Biomedical Research Centre as an Interdisciplinary Research Award. Ethical approval has been requested. Findings are expected to be produced by June 2025, and will be disseminated at symposia, conferences and in journal publications.

**Strengths and limitations of this study:** *Strengths:*

- This work highlights the importance of investigating the use of additional clinical variables to those used in NEWS2, in the development of a new early warning score
- The study design was informed by an evidence synthesis of the literature

*Limitations:*

- Some retrospective data sets may be of low quality and/or incomplete
- External validation will be needed to test algorithm generalisability

## 1. Introduction

The second National Early Warning Score (NEWS2) system is a widely used tool for the systematic documentation and identification of clinical deterioration. It has been demonstrated to be effective at reducing in-hospital mortality [1–4], it facilitates effective communication between clinicians and enables timely interventions to improve patient outcomes, but NEWS2 has limited positive and negative predictive accuracy [5–7], particularly in predicting adverse events beyond 24 hours [8–11].

The use of digital technologies in healthcare presents an opportunity to evaluate whether the inclusion of additional routinely collected variables would improve the predictive accuracy of an early warning score and thus reduce patient risk. The same technologies could then be used to provide clinical decision support using artificial intelligence-derived algorithms that would not be practical using paper observation charts and healthcare records [1,11].

This further personalisation of risk prediction is increasingly important in our ageing population. Older adults are particularly vulnerable to sudden changes in physiological status [13], but this group was not included in the development of NEWS2 [14]. Improved predictive accuracy in this growing frail and vulnerable group could significantly improve patient outcomes [14–16].

In an evaluation of the performance of the original NEWS, it demonstrated consistent predictive accuracy across multiple patient groups [5]. However, the performance of NEWS2 varies between care settings, and between patients with different characteristics [11,17,18]. NEWS2 has been found to have significantly lower specificity, sensitivity, and positive predictive value for those who were at risk of type II respiratory failure (T2RF), but higher specificity and positive predictive value for those with documented T2RF status, then the original NEWS [17]. The accuracy of NEWS has also been found to vary between care settings [18]. These observations suggest that the inclusion of individualised patient and care setting characteristics may improve the predictive accuracy, sensitivity and specificity of the system and thus improve outcomes in a range healthcare settings and in diverse patient groups.

To determine which variables may improve the risk prediction of NEWS2 we have reviewed six databases (CINAHL, PubMed, Embase, ScienceDirect, Cochrane Library and Web of Science) [19]. Our preliminary findings show the demographic variables including age and ethnicity [6,20], trend data [21–23], and adjustments to component weighting [12,24] may improve on the performance of NEWS2 in predicting clinical deterioration.

The principal aim of this work is to develop an algorithm that can improve on the predictive accuracy and thus clinical value of the NEWS2 system. The objectives of this study are:

1. To determine which additional variables have been previously used to enhance the accuracy of clinical early warning scores, including NEWS2;
2. To collect a large set of patient data that links observations, demographic, and admissions data with key deterioration-related outcomes;
3. To use these data to train an artificial intelligence (AI) model to determine which variables, and what weightings, provide optimal predictive accuracy;
4. To test the accuracy, sensitivity, and specificity of the algorithm;
5. To consider how the new model could be applied across a wide range of healthcare settings, including different physiological observation equipment, different electronic patient record systems, or in paper-based clinical record systems.

## 2. Methods and Analysis

The study is a retrospective cohort study using approximately 7 years (2017-2023) of historical, anonymised patient data from the Newcastle upon Tyne Hospitals NHS Foundation Trust to explore ways in which a new alerting system could improve on NEWS2. We will recruit PPIE members to join the project steering committee through well-established PPIE processes in the Newcastle BRC Informatics and Precision Care for an Ageing Population theme and Newcastle University’s Clinical Trials Unit. PPIE members will be included in all project management group meetings and will provide their lived experience with hospital monitoring to highlight any concerns or barriers they foresee in the adoption and implementation of a modified NEWS system. We will recruit at least one older adult to ensure that we are including a perspective that has been missing in the development of previous NEWS systems.

### Data collection

We will use a large number of routinely collected patient variables linked with individual patient outcomes and use these linked data to determine which variables, and with which algorithm, best predicts outcomes. The Newcastle Hospitals NHS Foundation Trust has an established information governance system for the use of anonymised patient data for research, which has been successfully used in the ADMISSION [32] and AI-Multiply [33] studies. A data scientist at the Trust will exclude all data from patients who have opted out of their data being used for research purposes through local or national processes. The data will then be de-identified before the research team can access it within the Trust’s IT network for training and testing the algorithm.

**Figure 1.**
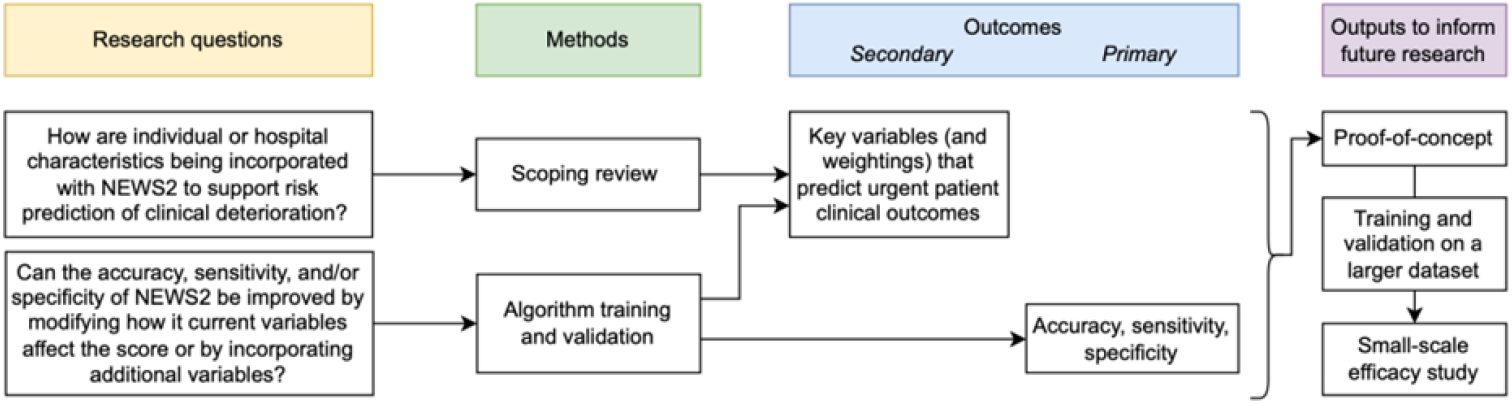
Study logic diagram

### Data analysis

Deidentified data from the Trust’s clinical data marts, including patient observations, admissions information, demographic data, and outcomes data, will be divided into training and testing subsets. Using these data sets, we will train and test an algorithm that optimises the variables and their weightings to predict the risk of key clinical outcomes, including mortality, ITU admission, sepsis and cardiac arrest, to demonstrate a proof of concept for a modified scoring system. By the end of this study in April 2025, we will have trained and tested an algorithm based on 7 years of historical patient data. We will then test the new system on data in other NHS Trusts across the UK to assess its generalisability. In 2026, we will conduct implementation studies in Newcastle and then in other NHS Trusts to assess the adoption of the system and gather initial data on patient outcomes to construct a learning healthcare system to evaluate real-world performance of the algorithm and facilitate continual improvement. By 2029, we aim to implement the system in NHS Trusts across the UK and conduct large-scale trials to evaluate its effectiveness at improving patient outcomes.

### Intended outputs

The aim of this project is to improve on the predictive accuracy of the NEWS2 scoring system, particularly its accuracy over more than 24-hours, and its predictive value in older patients and children. It will investigate whether using the currently collected data differently and the inclusion of additional data would result in an improved algorithm. There are two main ways in which this could improve patient outcomes: 1) Improving the system’s ability to accurately predict patients who will deteriorate within and beyond 24-hours would give healthcare professionals the opportunity to optimise their care and prevent deterioration, and 2) reducing false positive alerts would allow care to be concentrated where it will achieve the greatest benefit.

To be of national and international importance, the tool must meet the needs of patients throughout their life-course. It is clearly important to examine the predictive accuracy of any new early warning system in older people, but there is also limited outcome data for use of such systems in children. On 3 November 2023, NHS England announced the roll-out of a national Paediatric Early Warning System (PEWS) [25], and we will collaborate with the PEWS team in Newcastle Hospitals to develop a new evidence-based clinical alerting system for deteriorating children.

The tool must also be usable in the widest possible range of healthcare settings. As many hospitals worldwide do not have a high degree of digital maturity [26,27] and rely on paper-based documentation [28,29], we aim to develop a scalable paper- or smart-phone application-based system that can achieve the highest possible predictive accuracy and clinical value from an improved algorithm [30].

The overall aim is to incorporate an improved early warning system in an integrated Learning Health System. This requires the technical capacity to routinely collect outcomes data and provide structured opportunities to test and improve the algorithm for a changing population. It also needs robust mechanisms for capturing and using clinical feedback on its implementation, successes, and barriers in a wide range of care contexts [31].

## 3. Ethics and Dissemination

An application for the ethical approval of this study has been submitted to the UK The Health Research Authority Integrated Research Approval System and is under consideration at the time of manuscript submission. The study findings are expected to be produced in April 2025 and will be disseminated symposia, conferences, clinical meetings and academic peer-reviewed publications. PPIE members will help guide the dissemination of the results of the study to the population through publicly accessible forums. Further funding applications will be prepared to continue this programme of work by applying the new algorithm to other Trusts across the UK and other hospital systems around the world, to test its generalisability and refine its performance. Patient data will be stored only on secure NHS Trust systems. Other data available to the research team will be stored securely on Newcastle University systems until the study is complete. Analyses generated by the study will be available upon publication for 10 years from the PI on reasonable request.

## 4. Conclusion

This work has identified potential areas of improvement to the NEWS2 system and outlined a project for improving on its ability to predict adverse outcomes through the use of additional patient data and/or changes to the scoring process. The project’s outputs will be shared widely to facilitate collaboration between healthcare professionals, researchers, and policymakers to support the implementation of improved clinical early warning systems in diverse populations.

## Data Availability

All data produced in the present study are available upon reasonable request to the authors.

## Abbreviations

NEWS: National Early Warning Score

## CRediT author statement

**Chris Pummer:** Conceptualisation, Funding acquisition, Writing - review & editing

**Madison Milne-Ives:** Methodology, Writing - Original Draft

**Cen Cong:** Writing - Original Draft, Writing - Review & Editing

**Lynsey Threlfall:** Writing - review & editing

**Edward Meinert:** Conceptualisation, Supervision, Writing - review & editing

